# Chronic treatment with hydroxychloroquine and SARS-CoV-2 infection

**DOI:** 10.1101/2020.06.26.20056507

**Authors:** A Ferreira, A Oliveira-e-Silva, P Bettencourt

## Abstract

**Background:** Hydroxychloroquine sulphate (HCQ) is being scrutinized for repositioning in the treatment and prevention of SARS-Cov-2 infection. This antimalarial drug is also chronically used to treat patients with autoimmune diseases.

**Methods:** By analyzing the Portuguese anonymized data on private and public based medical prescriptions we have identified all cases chronically receiving HCQ for the management of diseases such as systemic lupus erythematosus, rheumatoid arthritis, and other autoimmune diseases. Additionally, we have detected all laboratory confirmed cases of SARS-CoV-2 infection and all laboratory confirmed negative cases in the Portuguese population (mandatorily registered in a centrally managed database). Cross linking the two sets of data has allowed us to compare the proportion of HCQ chronic treatment (at least 2 grams per month) in laboratory confirmed cases of SARS-CoV-2 infection with laboratory confirmed negative cases.

**Results:** Out of 26,815 SARS-CoV-2 positive patients, 77 (0.29%) were chronically treated with HCQ, while 1,215 (0.36%) out of 333,489 negative patients were receiving it chronically (P=0.04). After adjustment for age, sex, and chronic treatment with corticosteroids and/or immunosuppressants, the odds ratio of SARS-CoV-2 infection for chronic treatment with HCQ has been 0.51 (0.37-0.70).

**Conclusions:** Our data suggest that chronic treatment with HCQ confer protection against SARS-CoV-2 infection.

## Introduction

Several *in vitro* studies have shown chloroquine phosphate and hydroxychloroquine sulphate (HCQ) to be effective in both preventing and treating SARS-CoV-2 infection in isolated cells (1-3). Chloroquine phosphate and HCQ were also found effective in the treatment of COVID-19 patients in small nonrandomized or unblinded clinical studies (4-6). A recent observational study has shown no association between HCQ use and the composite endpoint of intubation or death in hospitalized patients (7). However, up until today there are no available results from well designed, randomized, double blinded, placebo controlled clinical trials showing any evidence of a therapeutic or preventive effect of these drugs in SARS-CoV-2 infection.

These two drugs are also chronically used to treat systemic lupus erythematosus (SLE), rheumatoid arthritis (RA) and other autoimmune diseases. SLE patients are at increased risk of viral infections (8). Increased ratios of serious bacterial and viral infections have been shown or are expected in different autoimmune diseases (8-11). Thus, patients with these diseases should be expected to have increased ratios of SARS-CoV-2 infection as compared to the general population. Antimalarials appear to have a protective effect against infections in patients with SLE (11, 12).

## Objectives

To evaluate the effect of chronic treatment with HCQ for other medical conditions in SARS-COV-2 infection incidence.

## Patients and methods

### Data collection

The Portuguese National Health Service (Serviço Nacional de Saúde – SNS) has a centrally controlled electronic database where all drug prescriptions, both from public and private medical care, are systematically registered (Prescrição Electrónica de Medicamentos – PEM, Serviços Partilhados do Ministério da Saúde – SPMS). In Portugal, all prescriptions are mandatorily made through a personal computer platform (there are no prescriptions on paper), in both public and private health services and are recorded in that centralized database.

A specific database allows the registry of obligatory notifiable diseases, including the suspected and confirmed cases of SARS-CoV-2 infection (Sistema Nacional de Vigilância Epidemiológica – SINAVE MED, Direcção Geral da Saúde – DGS). Another database allows the registry of all polymerase chain reaction (PCR) tests for SARS-CoV-2 performed in the country (SINAVE LAB, DGS).

Anonymized data were extracted from these databases. By analyzing these sets of data, we were able to detect all patients with SARS-CoV-2 confirmed infections and all clinically suspected but non-confirmed patients between Mars 2, 2020 (the date of the first Portuguese case) and the moment of the analysis. All cases prescribed with HCQ between January 1, 2019 and December 31, 2019 for other medical conditions were documented too. Additionally, we identified all prescriptions of corticosteroids (prednisolone, methylprednisolone, dexamethasone and deflazacort) and/or immunosuppressants (cyclophosphamide, azathioprine, cyclosporine, methotrexate, or mycophenolate mofetil) in the same period.

### Case definitions

Suspected case: patient with acute respiratory infection (sudden onset of fever or cough or respiratory distress) with no other aetiology explaining the condition and with a history of travel or residence in areas with active community transmission of SARS-CoV-2 within 14 days prior to onset of symptoms; or patient with acute respiratory infection and a history of contact with a confirmed or likely case of infection with SARS-CoV-2 or CoVID-19 within 14 days prior to the onset of symptoms; or patient with severe acute respiratory infection, requiring hospitalization with no other aetiology.

Confirmed case: patient with laboratory confirmation of SARS-CoV-2 infection by nasopharynx swab PCR test, regardless of signs and symptoms.

Since there were many unidentified records in SINAVE LAB (including expatriates), all cases in which at least one positive PCR test could be detected in the laboratory database were considered positive; all the other cases were checked against the SINAVE MED database and considered positive if registered as a confirmed case in this database (according to case definition) or negative otherwise.

To be considered under chronic treatment with HCQ, cases had to be prescribed with at least 2 grams of HCQ per month, on average. Chloroquine phosphate is not available in Portugal.

Patients with, at least, 6 months on corticosteroids and/or immunosuppressants were considered under chronic immunosuppressant treatment.

For each case, the first 2019 HCQ prescription was identified (index prescription). Then, the total HCQ dose (in grams) prescribed from the index prescription (inclusive) was quantified up until the last day of 2019. Finally, the average monthly dose of HCQ was computed. The same methodology was used for corticosteroids and immunosuppressants, but the number of prescriptions (not grams) was quantified.

Non-residents in Portugal were excluded from the study.

### Data Extraction

Anonymized raw data were extracted from the abovementioned databases on July 2, 2020. The raw data were revised to ensure internal consistency and transformed into a structured database for analysis.

The SINAVE LAB database contained the registry of 679,265 PCR tests carried out in Portugal up to the moment of the extraction. Of these, 189,734 were not identifiable and were excluded from the analysis. However, by cross-referencing the information from SINAVE LAB with SINAVE MED, it was possible to identify 360,552 suspected cases of SARS-CoV-2 infection.

The extraction from PEM database contained the registry of 26.735 patients who were prescribed the study medications between January 1, 2019 and December 31, 2019, including the number of prescriptions and their dates.

### Data analysis

We have analyzed the prevalence of HCQ chronic treatment in all PCR tested patients, comparing PCR positive cases with PCR negative cases.

### Statistical analysis

Numeric variables are presented as mean ± standard deviation. Proportions are presented as percentages.

Student’s t-test was used to test for differences between numeric variables. The chi-square test was used to test for differences between categorical variables.

Logistic regression was used to adjust the effect of the independent variable to confounding factors, like sex, age, and immunosuppressive treatment.

### Ethics

This study was approved by the Ethics Review Board of Centro Hospitalar Universitário de São João and Faculdade de Medicina da Universidade do Porto.

## Results

The first case of SARS-CoV-2 infection in Portugal was recorded on 2 March 2020. Since then, until the moment we analyzed the data, 360,304 patients with suspected SARS-CoV-2 infection had a final case definition (positive PCR test or negative PCR test). In 248 cases the PCR test result was inconclusive and had not yet been repeated at the time of data extraction. Of these 360,304 cases, 26,815 were confirmed by a positive PCR test. The rest had negative PCR tests.

In the set of patients with case definition, 1,292 received HCQ (at least 2 grams per month) and 897 were prescribed with corticosteroids and/or immunosuppressants for 6 months at least.

### Univariate analysis

The results of univariate analysis are summarized in table 1. SARS-CoV-2 positive patients were older and more frequently of the male gender than negative patients.

**Table 1.** Demographic characteristics, HCQ and corticosteroid and/or immunosuppressive treatment in SARS-CoV-2 positive and negative cases and statistical significance for their differences (P) in univariate analysis.

|  | SARS-CoV-2 Positive | SARS-CoV-2 Negative | P |
| --- | --- | --- | --- |
|  | n = 26,815 | n = 333,489 |  |
| HCQ | 77 (0.29%) | 1,215 (0.36%) | 0.04 |
| Male gender | 11.122 (41.5%) | 129.289 (38.9%) | <0.0001 |
| Age (years) | 52.2±22.4 | 50.8±22.4 | <0.0001 |
| CCT/Immunos. | 82 (0.31%) | 81 (0.24%) | 0.05 |

CCT – corticosteroids; Immunos. – immunosuppressants (treatment with corticosteroids – prednisolone, methylprednisolone, deflazacort or dexamethasone, and/or immunosuppressants – cyclophosphamide, cyclosporine, methotrexate, azathioprine, or mycophenolate mofetil for at least 6 months).
HCQ – at least 2 grams per month, on average.
Age was missing in 776 cases (2 positives and 774 negatives). Gender was missing in 822 cases (1 positive and 821 negatives). The results exclude missing data.

The proportion of HCQ chronic treatment was higher in negative patients (0.36% vs. 0.29%, P = 0.04) and the inverse was true for corticosteroids and/or immunosuppressants chronic treatment (0.24% vs. 0.31%, P = 0.05). The unadjusted odds ratio of a positive test for HCQ chronic treatment was 0.79 (0.63-0.99).

The incidence of SARS-CoV-2 infection in patients receiving HCQ was 5.96%, while in the other patients it was 7.45%

### Multivariate analysis

As shown in table 2, after adjustment for demographic characteristics and immunosuppressive treatment, HCQ remained independently and negatively associated with SARS-CoV-2 infection. The adjusted odds ratio of a positive PCR test for HCQ chronic treatment was 0.51 (0.37-0.70). Immunosuppressive treatment was found to be positively associated with SARS-CoV-2 infection with an odds ratio of 2.06 (1.51-2.82).

**Table 2.** Odds ratios (and 95% CI) of SARS-CoV-2 infection for study variables in multivariate analysis (logistic regression)

|  | OR | 95%CI |
| --- | --- | --- |
| HCQ | 0.51 | 0.37-0.70 |
| Male gender | 1.11 | 1.09-1.14 |
| Age (20-year increment) | 1.06 | 1.05-1.07 |
| CCT/Immunos. | 2.06 | 1.51-2.82 |

CCT – corticosteroids; Immunos. – immunosuppressants (treatment with corticosteroids – prednisolone, methylprednisolone, deflazacort or dexamethasone and/or immunosuppressants – cyclophosphamide, cyclosporine, methotrexate, azathioprine, or mycophenolate mofetil for at least 6 months).
HCQ – at least 2 grams per month, on average.
The logistic regression model included 359,476 cases (positive cases – 26,813; negative cases – 332,663) due to missing data.

## Discussion

As far as we know, this is the first study retrospectively assessing the relationship between chronic treatment with HCQ and SARS-CoV-2 infection in a large sample of patients at a nation-while level.

In this study, we only included patients chronically prescribed with HCQ during 2019, whose prescriptions continued into 2020. This way, we were able to exclude those who have started taking HCQ aiming to treat or prevent SARS-CoV-2 infection. Otherwise, this would be a confounding factor in the analysis of the data.

Although we have no information on comorbidities of the patients registered in the databases used in this study, it seems fair to assume that patients who chronically received HCQ at a dose of at least 2 grams per month have SLE, RA or other autoimmune diseases. In fact, in Portugal, HCQ is approved for the treatment of SLE, discoid lupus erythematosus, RA, juvenile idiopathic arthritis, and for the prophylaxis and treatment of malaria, which is not endemic (only imported cases). In the last case, the dose and duration of the treatment are much lower than the ones selected for inclusion in this study. Thus, patients using HCQ for prophylaxis or treatment of malaria were excluded from the study by the inclusion criteria we have used. The prevalence of SLE and RA in Portugal is 0.1% and 0.7%, respectively (13). These figures have allowed us to assume that chronic treatment with HCQ in such doses is a surrogate indicator of an autoimmune disease diagnosis.

Patients with SLE or RA have numerous cellular and humoral abnormalities that contribute to an increased susceptibility to infectious agents (14-16). SLE patients have a 3.9 times higher incidence rate of severe herpes simplex virus infection than controls (9). SLE patients treated with glucocorticoids alone also have a 3.9-fold hazard ratio for severe infections when compared to patients treated with HCQ alone (11). The prevalence of papillomavirus and cytomegalovirus is supposedly higher in patients with autoimmune diseases too (9,10). Besides the negative effect of the immunologic disturbance, these patients usually receive immunosuppressive drugs too, making them even more susceptible to infection (14-16).

Thus, the available data suggest that autoimmune patients are at increased risk of viral infections. The benefits of HCQ treatment in this context, if any, depend on its ability to prevent those infections. The COVID-19 risk scoring guide of the British Society for Rheumatology (17) seems to adopt the same orientation, by scoring with zero the use of HCQ while it scores with three high-dose corticosteroids and cyclophosphamide. Our data give further support to this suggestion.

We were able to show that patients taking HCQ have had reduced odds of SARS-CoV-2 infection. This effect was most evident after adjustment for age, sex, and chronic immunosuppressive treatment, increasing the likelihood that chronic treatment with HCQ actually has a protective effect against SARS-CoV- 2 infection.

A recent paper compared the proportions of HCQ use in SARS-CoV-2 positive and negative patients (18), finding no differences between the two groups. However, as study limitation, the authors stated that the duration of treatment was not documented and only 3 patients received HCQ in the positive cases group. An Indian study showed that HCQ (in the dosage currently used in the prophylaxis of malaria and after four maintenance doses) was effective in preventing SARS-CoV-2 infection in health care workers (19), while another paper showed no statistically significant effect of HCQ (in the dose of 600 mg/day for four days, after a loading dose of 800 mg plus 600 mg after 6 to 8 hours) in postexposure prevention (20). The first two studies evaluated the effectiveness of HCQ as pre-exposure prophylaxis, while the latter tested its effectiveness as post-exposure prophylaxis. Taken together, these results suggest that a relatively long period of HCQ treatment may be necessary in patients without prior contact with the virus to obtain a preventive effect. We believe our data supports this hypothesis.

Available data show that there was a significant increase of HCQ prescriptions in March 2020 and in the first two weeks of April 2020. Additional studies are needed to understand whether this increase in HCQ consumption has had any effect on the evolution of the outbreak in Portugal and on the incidence of adverse events.

In conclusion, our findings suggest that HCQ might have a preventive effect of SARS-CoV-2 infection.

### Statistical limitations

Despite having a considerable number of lab tests that were missing in the official electronic registries, we believe that we have captured all suspected confirmed and unconfirmed cases of SARS-CoV-2 infection by checking the lab data (SINAVE LAB) against de the obligatory registry database (SINAVE MED). Furthermore, all positive and negative cases included in our sample were confirmed by PCR testing, giving consistency to our results.

The fact that we do not have information on comorbidities may be a major limitation of the study, because it did not allow us to exclude a potential selection bias – for example, physicians may have decided to prescribe HCQ to patients with fewer comorbidities, and because it was not possible to adjust the effect of HCQ to such comorbidities. Thus, the observed effect could be related to a better health status of patients and not to a preventive action of HCQ. However, clinical guidelines recommend the use of antimalarials in all cases (21), regardless of the existence of comorbidities (namely, those that may affect resistance to infection, such as chronic renal failure). It is therefore to be expected that physicians have followed those recommendations and that no such selection bias has occurred.

We do not have any data on the therapeutic compliance of patients, and this is a study limitation too.

Although the proportions of HCQ chronic treatment are low, the difference in proportions between the two groups is unlikely to be cancelled out by the study pitfalls mentioned above.

### Study implications

Considering the low toxicity (22-24) and the enormous clinical experience with HCQ utilization, these results strongly suggest it should continue to be used in patients with autoimmune diseases, mostly in the context of the SARS-CoV-2 pandemic. Whether it should be used as a pre-exposure prophylactic measure in other risk groups or in the general population is unknown. We believe that our findings suggest that at least it should be considered for this purpose. The lockdown is severely affecting the world’s economy, increasing unemployment, and inducing more suffering than the disease itself. Eventually, its effects on the welfare of the populations will be much worse than the potential adverse effects of HCQ prophylaxis. Furthermore, it cannot be maintained for an extended period. Thus, while waiting for an effective and safe vaccine, a chemoprophylaxis-based strategy, using this inexpensive and safe drug, continuously monitored and modified according to the results of ongoing clinical trials, may be justified.

## Data Availability

Databases with the raw data and a structured database for analysis will be available for the reviewing process.

